# Association of Item-Level Responses to Cognitive Function Index with Tau Pathology and Hippocampal volume in The A4 Study

**DOI:** 10.1101/2024.09.16.24313705

**Authors:** Idris Demirsoy, Elham Ghanbarian, Babak Khorsand, Bhargav T. Nallapu, Ali Ezzati

## Abstract

**Background:** Alzheimer’s Disease (AD) has a lengthy asymptomatic preclinical phase during which individuals may show pathological signs like β-amyloid (Aβ) pathology and tau tangles without noticeable objective cognitive impairments. Subjective cognitive impairment reports may offer valuable and early insights into individuals’ cognitive functioning and serve as indicators of early stages of cognitive decline.

**Objective:** To investigate the associations of the item-level response to Cognitive Function Index (CFI) by participant and study partner with tau pathology and adjusted hippocampal volume (HVa).

**Method:** Participants were 339 cognitively unimpaired, Aβ positive, individuals enrolled in the Anti-Amyloid Asymptomatic Alzheimer’s (A4) Study who underwent tau-PET imaging. Participants and their study partners assessed subjective changes in cognition and function over the past year using the 15-item CFI questionnaire. For each CFI item, the relationship among tau, HVa, and CFI reports was investigated.

**Result:** Participants were on average 72.38 (SD = 4.87) years old, 58.1% were female, and 23.6% were tau positive. Higher tau_MTL_ was significantly associated with participant report of decline on three CFI items including depending on written notes, seeing a doctor for memory concern, and feeling lost while navigating. Higher tau_MTL_ was associated with study partner report of decline on two different items: needing help from others to remember appointments/occasions and asking same questions. Additionally, HVa was linked to challenges with driving for participants and noticeable memory decline for study partners.

**Conclusion:** We showed that early changes reported on specific items of the CFI are associated with higher tau_MTL_ and lower HVa in Aβ+ participants. Different CFI items were associated with tau and hippocampal volume for participants and study partners, highlighting the importance of both perspective.

## Introduction

Alzheimer’s Disease (AD) is characterized by a lengthy asymptomatic preclinical phase, during which individuals who appear cognitively normal may already exhibit the pathological hallmarks of the disease, including beta-amyloid (Aβ) pathology and tau tangles in their brains [1]. Clinical trials for AD have been increasingly focused on these early disease stages. However, targeting these early stages poses challenges because most individuals in the preclinical phases do not show noticeable cognitive or functional impairments [2]. Therefore, sensitive clinical methods are needed to detect and monitor subtle changes in cognitive functioning related to disease pathology before objective clinical impairment becomes apparent. This would enable more accurate quantification of disease progression and treatment effects [3].

Subjective cognitive impairment (SCI) is commonly reported by older adults and their study partners (informants), including neuropsychologically normal individuals [4]. The utility of SCI reports is multifaceted: First, they can help identify individuals experiencing perceived declines in cognitive function before objective deficits are measurable. Second, these reports offer insights into participants’ own experiences of their cognitive abilities, potentially revealing issues undetected by traditional neuropsychological tests. Third, SCI has predictive value, correlating with an increased risk of developing mild cognitive impairment (MCI) or dementia [5]. Fourth, SCI reports establish a baseline for perceived cognitive function to track future changes. Fifth, they aid in identifying participants in the study who are in the preclinical stages of cognitive decline, facilitating research on early markers and mechanisms of cognitive disorders. Finally, engaging patients in self-reporting their cognitive and functional well-being enhances their involvement in their medical care in general, and more specifically can promote proactive management of cognitive health. Given these utilities of SCI, it is imperative to studying the relationship between SCI, AD pathology, and clinical outcomes in clinical trials.

Previous studies investigating the relationships between SCI, cognitive performance, and amyloid deposition have shown that the associations between cognition-SCI and amyloid-SCI were influenced by racial differences and self-reported and study partner-reported SCI remained congruent across all groups, regardless of variations in study partner types [6]. Amariglio et al. examined the sensitivity of the SCI in tracking longitudinal cognitive changes among older adults without cognitive impairment at baseline [7]. They found that both SCI self-reported and SCI study partner-reported scores were linked to longitudinal cognitive decline, with evidence suggesting that self-reported scores may be more accurate in the early stages. The SCI thus emerges as a brief, yet informative, potential outcome measure that offers insights into functional abilities at the earliest stages of the disease. Subsequent analysis by the same group identified the specific types of subjective cognitive changes reported by participants and their study partners through item-level analysis in amyloid-positive (Aβ+) participants. Their findings indicated that participants were more likely than study partners to report changes on most SCI items; however, specific SCI items were associated with elevated Aβ levels in both groups, highlighting the importance of considering both sources of information in clinical trials [8]. Recently, Jadick et al. investigated the associations between tau deposition and preclinical Alzheimer’s disease (AD), particularly focusing on those in the earliest stages of the disease [9]. They observed that both self-report and study partner reports of SCI were associated with tau in addition to Aβ, indicating their utility in early detection of impairment.

In this study, we used data from participants in the Anti-Amyloid Asymptomatic Alzheimer’s (A4) Study [10] to study the association between the SCI and AD biomarkers at cross-section among cognitively unimpaired, Aβ+ individuals. Perceived SCI by participant and informant is measured in A4 using the Cognitive Function Index (CFI) [7, 8, 11]. We explored the overall and item-level associations of CFI with tau pathology and adjusted hippocampal volume (HVa). Additionally, we examined the relationship between demographic variables and discrepancies between participants’ self-reports and those of their study partners to understand the impact of differing perspectives on these subjective reports.

## Methods

### Participants

Data were gathered from participants involved in the A4 Study, which was carried out at 67 locations across the United States, Australia, Japan, and Canada. Each site received approval from their Institutional Review Board. Prior to participating, all participants gave their written consent. The A4/LEARN Study was overseen by the Alzheimer’s Therapeutic Research Institute (ATRI) at the University of Southern California, and the data were made accessible via the Laboratory for Neuro Imaging at the same university [12].

Initially, 6,763 cognitively normal individuals aged between 65 and 85 years were screened for enrollment in the A4 Study. Individuals who underwent screening for the A4 study were determined to be cognitively unimpaired, meeting criteria including a global Clinical Dementia Rating (CDR) score of 0, Mini-Mental State Exam (MMSE) score between 25 and 30, and a Logical Memory II subscale delayed paragraph recall (LM-IIa) score on the Wechsler Memory Scale-Revised (WMS-R) ranging from 6 to 18 [13]. Participants with a high brain amyloid burden, as measured by PET scans in key regions, qualified for the A4 study. Those who met all other criteria but exhibited lower amyloid levels were enrolled in the LEARN study, forming a separate group for the further investigation of their cognitive, clinical, and biological changes.

Following these criteria, 4,486 participants underwent ^18^F-florbetapir PET imaging [11, 14]. In a subgroup of participants, ^18^F-flortaucipir tau PET imaging was conducted. Amyloid positive individuals who also had measurements of PET tau were eligible for this study (N=339) (Figure 1).

**Figure 1.**
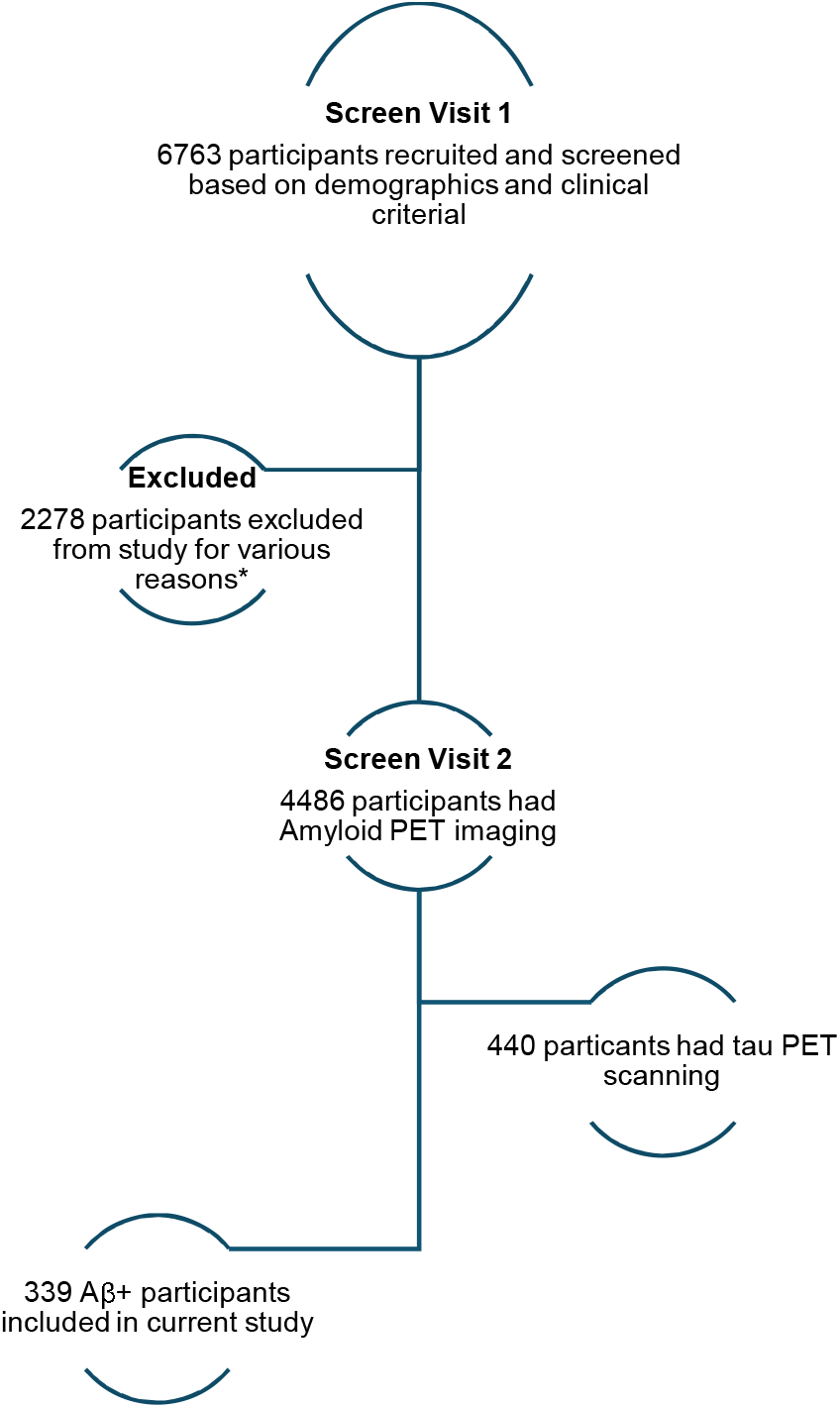
Flow chart of study participants.

### Cognitive Function Index

The Alzheimer’s Disease Cooperative Study (ADCS) designed the CFI to monitor variations in cognitive function over a one-year period [7, 15, 16]. Participants were required to possess sufficient literacy in one of the English, Japanese, or Spanish languages, as well as adequate hearing and vision abilities to complete the cognitive assessments. Additionally, a study partner was needed to give supplementary details regarding the participant’s cognitive functioning in daily life. These study partners were expected to have regular contact at least once a week with participants either by phone, in person, or via email. The CFI questionnaire is provided in two versions: one for participants to self-report their cognitive abilities, and another for study partners to report on the cognitive abilities of the participants [8]. Both participants and study partners receive the CFI by mail four weeks before each yearly evaluation. They are instructed to complete the CFI independently, with the stipulation that study partners may not consult with the participant but may consult with others.

The CFI consists of 14 questions, with identical questions for both the participant and study partner versions [7]. An additional question was added for the A4 study “In the past year, have you seen a doctor about memory concerns?” with the answer choices being Yes (1) or No (0) [7]. The total CFI score is calculated by adding up each item from the participant and study partner versions of the questionnaire. When calculating the CFI score, Amariglio et al employed a scoring system where Yes (2), Maybe (1), and No (0) [8]. Another study by Li et al have utilized a different scoring system [17]. Nevertheless, a higher CFI score indicates a greater degree of subjective cognitive complaints. Questions about driving, finances, and work performance include a “not applicable” response option. For these questions, from the available responses the the average was computed and used when the “not applicable” is selected [14].

### Amyloid PET Imaging and Determination of Aβ Status

^18^F-florbetapir PET imaging was conducted 50-70 minutes after administering 10 mCi of F-florbetapir [18]. Amyloid burden (classified as elevated (Aβ+) and eligible to proceed to screening or not elevated (Aβ-) and ineligible) was determined using an algorithm that combined quantitative standardized uptake value ratio (SUVr) methodology and qualitative visual interpretation performed at a central laboratory. An average cortical standardized uptake value ratio (SUVr), utilizing a whole cerebellar reference region of <1.15, was employed as the primary criterion to define not elevated amyloid (Aβ-). This quantitative approach was deemed to be more sensitive in detecting early amyloidosis in the preclinical stage of AD [8].

### Tau PET Imaging and Determination of Tau Status

Some of the participants in the A4 cohort underwent ^18^F-flortaucipir PET imaging, with image acquisition taking place between 90- and 110-minutes post-injection. For all brain regions standardized uptake value ratios (SUVRs) were calculated (tau SUVR) using the cerebellar gray matter as the reference tissue. The tau PET level was calculated for a region of interest (ROI): Medial Temporal Lobe (MTL) (unweighted average of bilateral entorhinal cortex and amygdala) [19]. The entorhinal cortex, despite being a smaller structure than the amygdala, has been shown to contribute more significantly in the early phases of tau accumulation [20], hence the MTL ROI is unweighted. The MTL ROI was adapted from a previously documented temporal meta-ROI, which is grounded in the widely recognized pattern of tau pathology progression from the MTL to the lateral temporal cortex [21, 22]. Thresholds for defining tau status (positive vs negative) in the ROI was set independently, based on the mean plus two standard deviations of tau levels observed in all Aβ-negative individuals within the A4 study [19]. The cut-off was determined to be 1.30 for tau_MTL_.

### Volumetric Magnetic Resonance Imaging (MRI)

All participants underwent volumetric segmentation of various cortical and subcortical brain regions using FreeSurfer 6.0. High-resolution three-dimensional (3D) T1-weighted MRI scans were processed, and quality control checks were performed with NeuroQuant (CorTechs Labs, San Diego, California), an automated segmentation tool cleared by the FDA for clinical application [12]. In this study, the original hippocampal volume (HV) was utilized as an indicator of neurodegeneration (N). To account for inter-personal variations in intracranial volume (ICV), the HVa was adjusted using residual-based correction via the following equation:

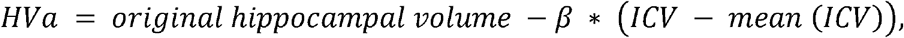

where β was obtained from regression equation between original HV and ICV.

### Genetic Measures

The most important genetic risk factor for AD is apolipoprotein E (APOE) ε4 allele. People who have one or two copies of the ε4 allele are much more likely to acquire AD pathology and its clinical symptoms than people who do not have this gene [18]. APOE ε4 status—0, 1, or 2 ε4 alleles— was included in our models as a categorical variable to account for its impact on outcomes.

### Statistical Analyses

Demographic information from participants and their study partners were summarized for the entire sample. Continuous data were described using mean and standard deviations and group differences were assessed using independent sample t-tests. For categorical data, analyses were conducted using chi square test and the Fisher’s Exact test. The Pearson correlation coefficients to evaluate the relationships among tau_MTL_, Aβ, and HVa. Multiple linear regression was used to assess the relationship between the total CFI scores (reported by participants or study partners), pathological tau levels (measured by PET imaging), and hippocampal volume (as a marker of neurodegeneration), adjusting for demographic variables.

Additionally, to investigate differences in endorsements on individual CFI items with imaging biomarkers variables for Aβ+ participants a series of logistic regression models were employed [23]. Models utilized dichotomous outcomes (0 = No endorsement, 1 = Yes/Maybe endorsement) and incorporated demographic, neuroanatomical, and pathological variables as predictors. A revised scoring method was implemented to account for missing data, where “not applicable” responses from participants were excluded from their overall scores. In such cases, the total score for each individual is calculated by summing the scores of the relevant items each participant answered and dividing by the total number of items for which they provided a response. This resulted in a final score between 0 and 1, which reflects the participants’ endorsement of cognitive concerns relative to the maximum achievable score based on their completed items. Finally, McNemar’s test was performed to assess the significant concordance in item level response patterns between patients and their study partners. All analyses were performed using R version 4.3.1 [24].

## Results

### Demographics

A total of 339 individuals Aβ+ individuals were eligible for this study. Among these participants, 23.6% were classified as tau positive in the medial temporal lobe (T+), and 58.1% were female. The T+ group had higher rate of APOE ε4 alleles compared to T-group. No significant differences were observed between the T+ and T-groups in terms of sex, educational background, marital status, or retirement status (Table 1).

**Table 1.**
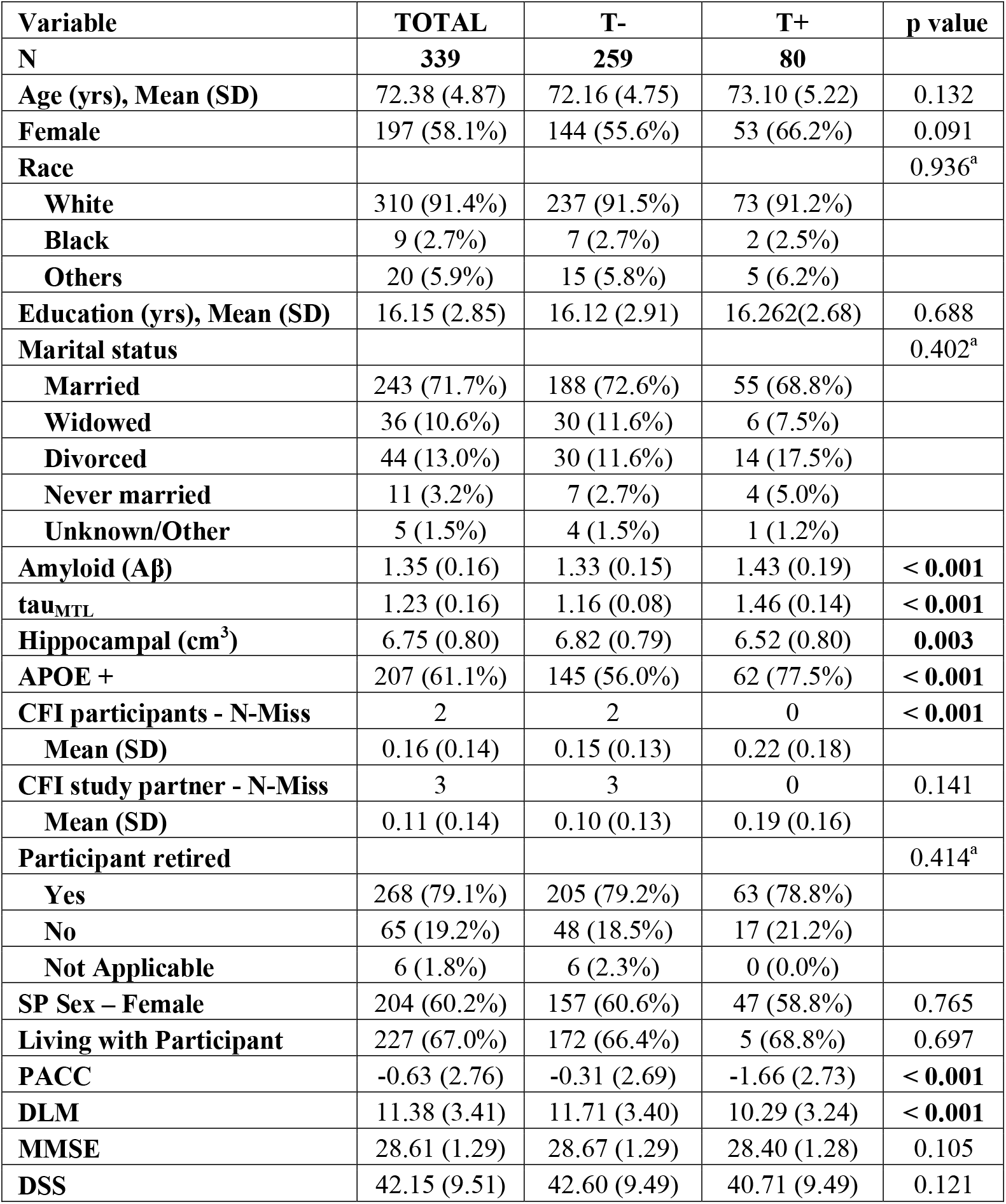
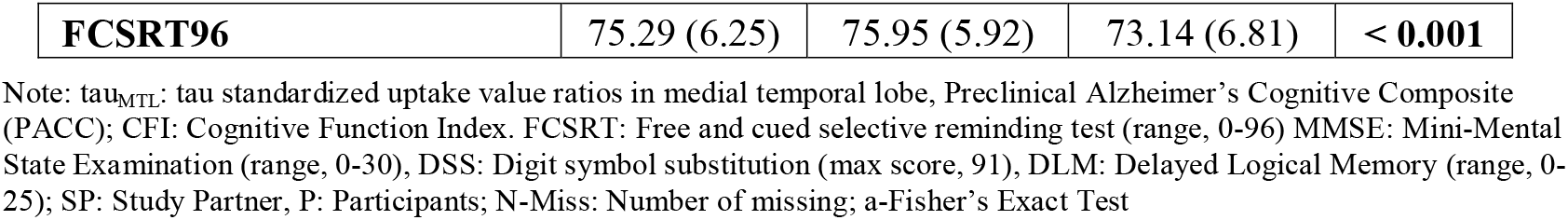
Amyloid Positive Participants stratified for tau MTL.

| **Variable** | **TOTAL** | **T-** | **T+** | **p value** |
| --- | --- | --- | --- | --- |
| **N** | **339** | **259** | **80** |  |
| **Age (yrs), Mean (SD)** | 72.38 (4.87) | 72.16 (4.75) | 73.10 (5.22) | 0.132 |
| **Female** | 197 (58.1%) | 144 (55.6%) | 53 (66.2%) | 0.091 |
| **Race** |  |  |  | 0.936<sup>a</sup> |
| **White** | 310 (91.4%) | 237 (91.5%) | 73 (91.2%) |  |
| **Black** | 9 (2.7%) | 7 (2.7%) | 2 (2.5%) |  |
| **Others** | 20 (5.9%) | 15 (5.8%) | 5 (6.2%) |  |
| **Education (yrs), Mean (SD)** | 16.15 (2.85) | 16.12 (2.91) | 16.262(2.68) | 0.688 |
| **Marital status** |  |  |  | 0.402<sup>a</sup> |
| **Married** | 243 (71.7%) | 188 (72.6%) | 55 (68.8%) |  |
| **Widowed** | 36 (10.6%) | 30 (11.6%) | 6 (7.5%) |  |
| **Divorced** | 44 (13.0%) | 30 (11.6%) | 14 (17.5%) |  |
| **Never married** | 11 (3.2%) | 7 (2.7%) | 4 (5.0%) |  |
| **Unknown/Other** | 5 (1.5%) | 4 (1.5%) | 1 (1.2%) |  |
| **Amyloid (Aβ)** | 1.35 (0.16) | 1.33 (0.15) | 1.43 (0.19) | **< 0.001** |
| **tau<sub>MTL</sub>** | 1.23 (0.16) | 1.16 (0.08) | 1.46 (0.14) | **< 0.001** |
| **Hippocampal (cm<sup>3</sup>)** | 6.75 (0.80) | 6.82 (0.79) | 6.52 (0.80) | **0.003** |
| **APOE +** | 207 (61.1%) | 145 (56.0%) | 62 (77.5%) | **< 0.001** |
| **CFI participants - N-Miss** | 2 | 2 | 0 | **< 0.001** |
| **Mean (SD)** | 0.16 (0.14) | 0.15 (0.13) | 0.22 (0.18) |  |
| **CFI study partner - N-Miss** | 3 | 3 | 0 | 0.141 |
| **Mean (SD)** | 0.11 (0.14) | 0.10 (0.13) | 0.19 (0.16) |  |
| **Participant retired** |  |  |  | 0.414<sup>a</sup> |
| **Yes** | 268 (79.1%) | 205 (79.2%) | 63 (78.8%) |  |
| **No** | 65 (19.2%) | 48 (18.5%) | 17 (21.2%) |  |
| **Not Applicable** | 6 (1.8%) | 6 (2.3%) | 0 (0.0%) |  |
| **SP Sex – Female** | 204 (60.2%) | 157 (60.6%) | 47 (58.8%) | 0.765 |
| **Living with Participant** | 227 (67.0%) | 172 (66.4%) | 5 (68.8%) | 0.697 |
| **PACC** | -0.63 (2.76) | -0.31 (2.69) | -1.66 (2.73) | **< 0.001** |
| **DLM** | 11.38 (3.41) | 11.71 (3.40) | 10.29 (3.24) | **< 0.001** |
| **MMSE** | 28.61 (1.29) | 28.67 (1.29) | 28.40 (1.28) | 0.105 |
| **DSS** | 42.15 (9.51) | 42.60 (9.49) | 40.71 (9.49) | 0.121 |
| **FCSRT96** | 75.29 (6.25) | 75.95 (5.92) | 73.14 (6.81) | **< 0.001** |

Note: tau<sub>MTL</sub>: tau standardized uptake value ratios in medial temporal lobe, Preclinical Alzheimer's Cognitive Composite (PACC); CFI: Cognitive Function Index. FCSRT: Free and cued selective reminding test (range, 0-96) MMSE: Mini-Mental State Examination (range, 0-30), DSS: Digit symbol substitution (max score, 91), DLM: Delayed Logical Memory (range, 0-25); SP: Study Partner, P: Participants; N-Miss: Number of missing; a-Fisher's Exact Test

We found a distinct tau pathology presence between the two groups, with the T+ group exhibited higher mean Aβ values (1.43, SD = 0.19) compared to the T-group (1.33, SD = 0.15) (p < 0.001). We also found a significant reduction in HVa in the T+ group (6.52, SD = 0.80, p = 0.003) (Table 1).

Compared to the T+, the T-group had higher cognitive performance across several objective cognitive measures, including the Preclinical Alzheimer Cognitive Composite (PACC), logical memory – delayed recall (DLM), and Free and Cued Selective Reminding Test (FCSRT96), all showing significant differences with p-values of < 0.001. The total CFI scores for participants were significantly higher in the T+ group (p < 0.001). However, the total CFI score reported by study-partner did not differ significantly between the two tau_MTL_ groups (p = 0.141). (Table 1).

### Total CFI score analysis

In models with participant-reported total CFI scores as the outcome (Table 2), higher tau_MTL_ was associated with the CFI scores (β = 0.15, p = 0.011). HVa was not associated with the total CFI scores in this model. The model examining the study partner-reported total CFI scores as the outcome showed a non-significant, positive trend for the association between tau_MTL_ and CFI scores (β = 0.11, p = 0.064). While participants were limited to Aβ+ individuals, for both participant and study partner CFI outcomes, higher Aβ levels were associated with higher CFI scores (Participant CFI: β = 0.12, p = 0.037; Study Partner CFI: β = 0.15, p = 0.008). (Table 2).

**Table 2.** Association of Total CFI Scores with Demographics and Imaging Biomarkers.

| Outcome | Participants reported CFI score |  |  | Study Partner reported CFI score |  |  |
| --- | --- | --- | --- | --- | --- | --- |
|  | Estimate | Std. Error | Pr(>\|t\|) | Estimate | Std. Error | Pr(>\|t\|) |
| (Intercept) | 0.02 | 0.12 | 0.887 | 0.15 | 0.11 | 0.195 |
| APOE+ | 0.07 | 0.12 | 0.566 | 0.02 | 0.12 | 0.869 |
| Female | -0.10 | 0.12 | 0.426 | **-0.27** | **0.12** | **0.026** |
| Age | 0.06 | 0.06 | 0.320 | -0.03 | 0.06 | 0.612 |
| Amyloid (Aβ) | **0.12** | **0.06** | **0.037** | **0.15** | **0.06** | **0.008** |
| *HVa* | -0.02 | 0.07 | 0.817 | -0.07 | 0.07 | 0.288 |
| tau<sub>MTL</sub> | **0.15** | **0.06** | **0.011** | 0.11 | 0.06 | 0.064 |

Note: tau<sub>MTL</sub>: tau standardized uptake value ratios in medial temporal lobe, HVa: adjusted hippocampal volume.

### Concordance between self-report and study partner assessment of CFI items

The responses to CFI items were relatively consistent between participants and their study partners, especially for items that received either high or low levels of endorsement. The items receiving the highest frequency of endorsements were d*ifficulty with remembering things* (Participants: 63%, Study Partners: 28%), *depending on written notes* (Participants: 46% vs Study Partners: 29%), and *misplacing things* (Participants: 32%, Study Partners: 24%). The items with the lowest frequency of endorsements included *struggling with appliances* (Participants: 3%, Study Partners: 4%), *struggling with financial tasks* (Participants: 4.9%, Study Partners: 3.7%), and *struggling with hobbies* (Participants: 6.9%, Study Partners: 3.6%) (Figure 2).

**Figure 2.**
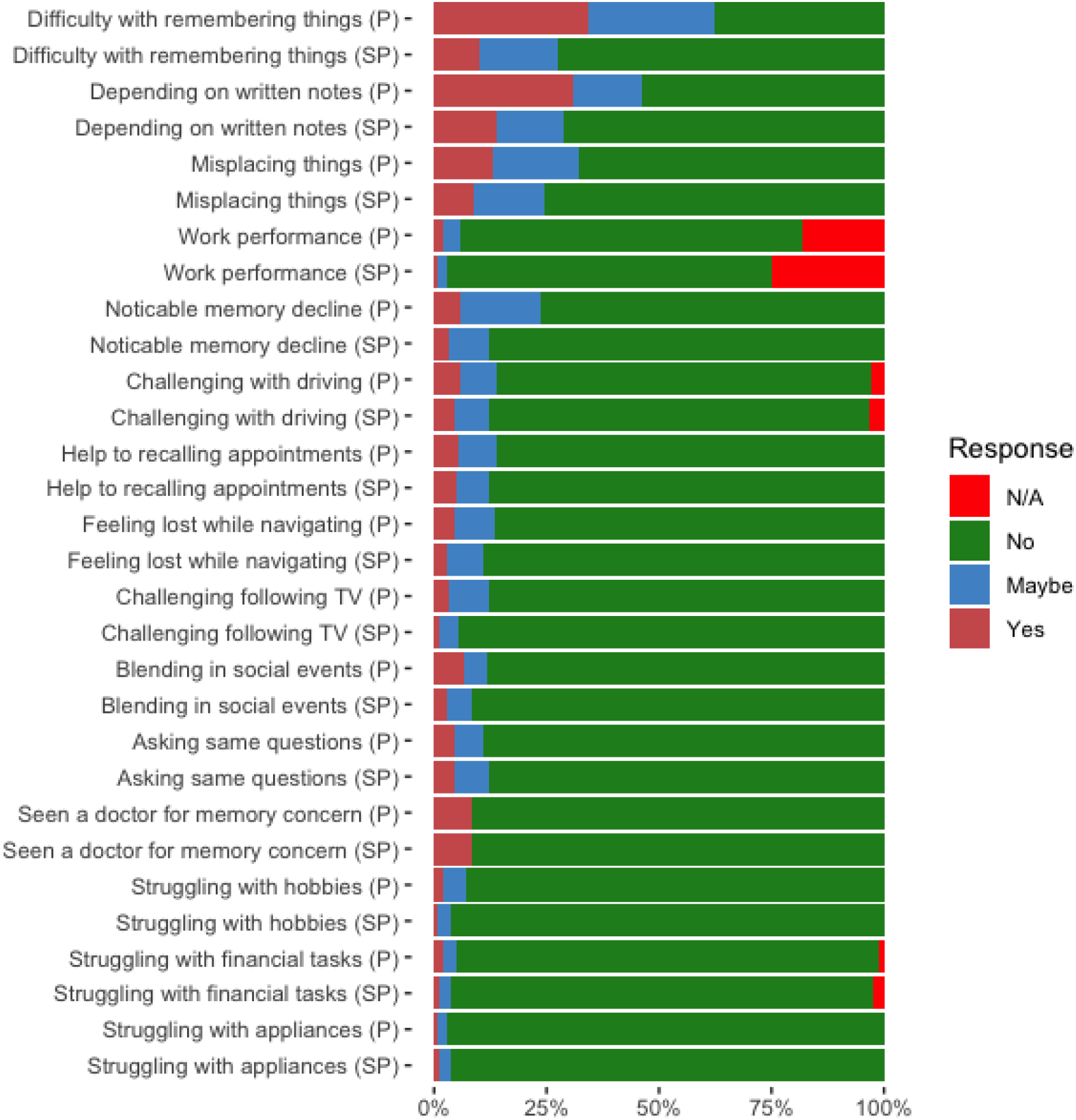
Proportion of endorsement on Cognitive Function Index (CFI) items as reported by participants and their study partners at the item-level analysis. (SP: Study Partner, P: Participants, N/A: Not applicable.)

### Item level analysis

We examined the association of item level CFI scores and ADRD biomarkers of amyloid, tau, and neurodegeneration (hippocampal volume). Three different models were developed as presented in Table 3. Model 1 explores the association between tau in the medial temporal lobe (tau_MTL_) and self-reported CFI item endorsement. Results indicated a significant association between four CFI items, participants reported *relying on written notes* (OR = 1.370, p = 0.011), *seeing a doctor for memory concerns* (OR = 1.743, p = 0.002), *feeling lost while navigating* (OR = 1.550, p = 0.003), and *work performance* (OR = 1.449, p = 0.046).

**Table 3.** Model comparisons for AB+ participants – tauMTL.

|  |  | Model 1 |  | Model 2 |  | Model 3 |  |
| --- | --- | --- | --- | --- | --- | --- | --- |
|  |  | Odds ratio | P value | Odds ratio | P value | Odds ratio | P value |
| Written | Aβ | 0.818 | 0.096 | 0.894 | 0.337 | 0.824 | 0.112 |
|  | tauMTL | **1.370** | **0.011** |  |  | **1.385** | **0.009** |
|  | HVa |  |  | 1.030 | 0.833 | 1.091 | 0.549 |
| Social | Aβ | **1.426** | **0.026** | **1.490** | **0.011** | **1.451** | **0.020** |
|  | tauMTL | 1.111 | 0.527 |  |  | 1.139 | 0.446 |
|  | HVa |  |  | 1.154 | 0.493 | 1.185 | 0.421 |
| Repeat | Aβ | 1.154 | 0.394 | 1.175 | 0.327 | 1.140 | 0.441 |
|  | tauMTL | 1.169 | 0.337 |  |  | 1.144 | 0.415 |
|  | HVa |  |  | 0.849 | 0.435 | 0.879 | 0.546 |
| Active | Aβ | 1.444 | 0.069 | 1.443 | 0.061 | 1.451 | 0.069 |
|  | tauMTL | 0.976 | 0.912 |  |  | 0.980 | 0.927 |
|  | HVa |  |  | 1.044 | 0.874 | 1.041 | 0.885 |
| Concern | Aβ | **1.574** | **0.016** | **1.734** | **0.003** | **1.546** | **0.022** |
|  | tauMTL | **1.743** | **0.002** |  |  | **1.692** | **0.004** |
|  | HVa |  |  | 0.699 | 0.177 | 0.833 | 0.495 |
| Recall | Aβ | 1.138 | 0.336 | 1.063 | 0.640 | 1.126 | 0.381 |
|  | tauMTL | 0.799 | 0.078 |  |  | 0.786 | 0.063 |
|  | HVa |  |  | 0.932 | 0.647 | 0.889 | 0.452 |
| Appliance | Aβ | 1.271 | 0.320 | 1.274 | 0.303 | 1.269 | 0.329 |
|  | tauMTL | 1.016 | 0.954 |  |  | 1.014 | 0.958 |
|  | HVa |  |  | 0.985 | 0.960 | 0.987 | 0.965 |
| Follow | Aβ | 1.309 | 0.083 | **1.419** | **0.020** | 1.331 | 0.069 |
|  | tauMTL | 1.269 | 0.114 |  |  | 1.291 | 0.097 |
|  | HVa |  |  | 1.078 | 0.709 | 1.138 | 0.524 |
| Lost | Aβ | 1.086 | 0.615 | 1.216 | 0.211 | 1.088 | 0.610 |
|  | tauMTL | **1.550** | **0.003** |  |  | **1.555** | **0.004** |
|  | HVa |  |  | 0.916 | 0.663 | 1.018 | 0.930 |
| Help | Aβ | **1.359** | **0.049** | **1.450** | **0.015** | **1.368** | **0.047** |
|  | tauMTL | 1.337 | 0.056 |  |  | 1.349 | 0.055 |
|  | HVa |  |  | 0.981 | 0.925 | 1.058 | 0.780 |
| Memory | Aβ | 1.008 | 0.949 | 1.069 | 0.593 | 1.006 | 0.961 |
|  | tauMTL | 1.278 | 0.051 |  |  | 1.274 | 0.056 |
|  | HVa |  |  | 0.938 | 0.678 | 0.981 | 0.900 |
| Misplace | Aβ | 1.237 | 0.085 | **1.299** | **0.030** | 1.232 | 0.093 |
|  | tauMTL | 1.266 | 0.051 |  |  | 1.259 | 0.060 |
|  | HVa |  |  | 0.917 | 0.557 | 0.957 | 0.771 |
| Money | Aβ | 1.367 | 0.190 | 1.375 | 0.173 | 1.340 | 0.226 |
|  | tauMTL | 1.149 | 0.581 |  |  | 1.120 | 0.656 |
|  | HVa |  |  | 0.825 | 0.547 | 0.848 | 0.609 |
| Drive | Aβ | 1.043 | 0.791 | 1.016 | 0.916 | 1.006 | 0.972 |
|  | tauMTL | 1.097 | 0.542 |  |  | 1.039 | 0.802 |
|  | HVa |  |  | **0.670** | **0.043** | 0.676 | 0.051 |
| Work | Aβ | 1.127 | 0.566 | 1.207 | 0.344 | 1.096 | 0.665 |
|  | tauMTL | **1.449** | **0.046** |  |  | 1.403 | 0.074 |
|  | HVa |  |  | 0.736 | 0.249 | 0.812 | 0.437 |

**Note:** tau<sub>MTL</sub>: tau standardized uptake value ratios in medial temporal lobe, HVa: adjusted hippocampal volume, Aβ : Beta-Amyloid. Model 1 involves tauMTL + Aβ , model 2 involves only HVa + Aβ and model 3 involves three of them. Note: all models involve APOE4, Sex, Age and Education.

Model 2 examined the association between HVa and item-level CFI endorsement. A specific CFI item demonstrated the notable associations with HVa. The item of participants *having challenges with driving* showed HVa as significant (OR = 0.670, p = 0.043).

In Model 3, the combined influence of tau_MTL_ and HVa on CFI items was evaluated (Figure 3). The findings remained consistent with those from the model 1. Specifically, participants endorsed three CFI items—*depending on written notes* (OR = 1.385, p = 0.009), having *seen a doctor for memory concerns* (OR = 1.692, p = 0.004), and *feeling lost while navigating* (OR = 1.555, p = 0.004).

**Figure 3.**
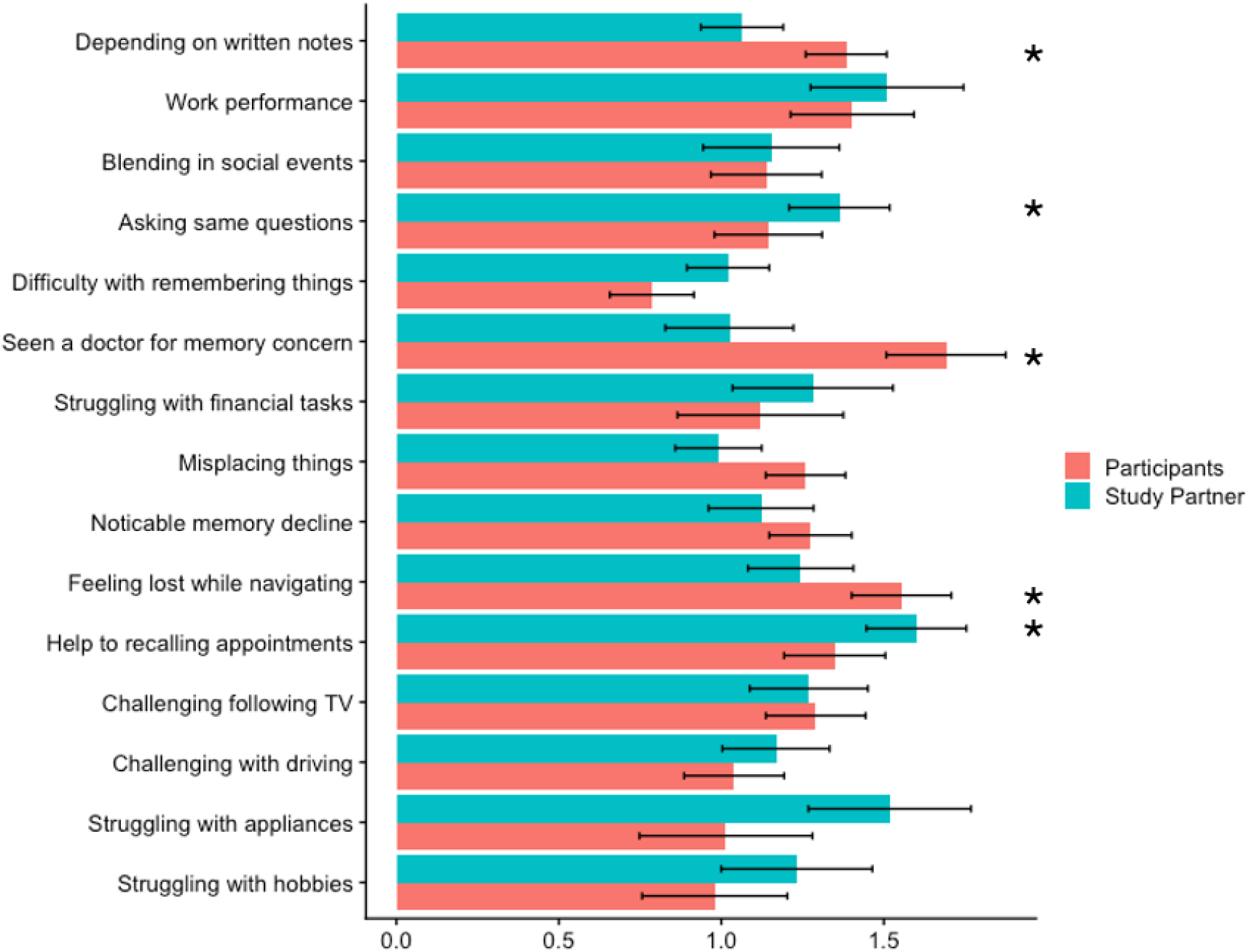
Odds of endorsement for CFI items among participants and their study partners according to Model 3. Asterisks indicate the significant p values for each item.

Next, we repeated the analysis for study partner-reported CFI items (Table 4). Model 1 explored the association between tau_MTL_ and study partner reported CFI item endorsement. The results showed that higher tau_MTL_ was only significantly associated with increased endorsement of one item: needing help recalling appointments (OR = 1.589, p = 0.002). Model 2 explored the association between HVa and study partner-reported CFI item endorsements. Smaller HVa was observed in those with higher endorsement in the *noticeable memory decline* item (OR = 1.589, p = 0.002). In model 3, which included both tau_MTL_ and HVa in the models, higher tau_MTL_ was seen in those with higher endorsement rate of two items: *asking same question* (OR = 1.363, p = 0.045), having *help the recalling appointment* (OR = 1.600, p = 0.002).

**Table 4.** Model comparisons for AB+ study partner – tauMTL.

|  |  | Model 1 |  | Model 2 |  | Model 3 |  |
| --- | --- | --- | --- | --- | --- | --- | --- |
|  |  | Odds ratio | P value | Odds ratio | P value | Odds ratio | P value |
| Written | Aβ | 1.091 | 0.492 | 1.092 | 0.477 | 1.076 | 0.569 |
|  | tau<sub>MTL</sub> | 1.084 | 0.520 |  |  | 1.064 | 0.625 |
|  | HVa |  |  | 0.850 | 0.290 | 0.860 | 0.331 |
| Social | Aβ | 1.259 | 0.282 | 1.294 | 0.220 | 1.246 | 0.315 |
|  | tau<sub>MTL</sub> | 1.164 | 0.462 |  |  | 1.154 | 0.495 |
|  | HVa |  |  | 0.904 | 0.721 | 0.936 | 0.815 |
| Repeat | Aβ | **1.411** | **0.025** | **1.542** | **0.004** | **1.439** | **0.019** |
|  | tau<sub>MTL</sub> | 1.330 | 0.059 |  |  | **1.363** | **0.045** |
|  | HVa |  |  | 1.100 | 0.636 | 1.182 | 0.412 |
| Active | Aβ | 1.145 | 0.577 | 1.163 | 0.519 | 1.092 | 0.725 |
|  | tau<sub>MTL</sub> | 1.297 | 0.257 |  |  | 1.232 | 0.369 |
|  | HVa |  |  | 0.646 | 0.176 | 0.682 | 0.238 |
| Concern | Aβ | 1.055 | 0.785 | 1.048 | 0.805 | 1.041 | 0.841 |
|  | tau<sub>MTL</sub> | 1.040 | 0.843 |  |  | 1.025 | 0.901 |
|  | HVa |  |  | 0.863 | 0.545 | 0.867 | 0.559 |
| Recall | Aβ | **1.527** | **0.001** | **1.536** | **0.001** | **1.529** | **0.001** |
|  | tau<sub>MTL</sub> | 1.019 | 0.879 |  |  | 1.021 | 0.869 |
|  | HVa |  |  | 1.011 | 0.944 | 1.015 | 0.925 |
| Appliance | Aβ | 1.016 | 0.956 | 1.144 | 0.615 | 1.028 | 0.921 |
|  | tau<sub>MTL</sub> | 1.498 | 0.098 |  |  | 1.518 | 0.095 |
|  | HVa |  |  | 0.993 | 0.984 | 1.094 | 0.794 |
| Follow | Aβ | 1.351 | 0.102 | 1.415 | 0.053 | 1.326 | 0.134 |
|  | tau<sub>MTL</sub> | 1.290 | 0.157 |  |  | 1.269 | 0.189 |
|  | HVa |  |  | 0.833 | 0.460 | 0.876 | 0.594 |
| Lost | Aβ | **1.382** | **0.041** | **1.423** | **0.022** | 1.353 | 0.059 |
|  | tau<sub>MTL</sub> | 1.293 | 0.109 |  |  | 1.244 | 0.178 |
|  | HVa |  |  | 0.729 | 0.133 | 0.767 | 0.213 |
| Help | Aβ | 1.130 | 0.439 | 1.269 | 0.114 | 1.136 | 0.423 |
|  | tau<sub>MTL</sub> | **1.589** | **0.002** |  |  | **1.600** | **0.002** |
|  | HVa |  |  | 0.952 | 0.802 | 1.052 | 0.797 |
| Memory | Aβ | **1.426** | **0.024** | **1.418** | **0.024** | **1.379** | **0.044** |
|  | tau<sub>MTL</sub> | 1.182 | 0.296 |  |  | 1.122 | 0.475 |
|  | HVa |  |  | **0.660** | **0.048** | 0.678 | 0.068 |
| Misplace | Aβ | 1.120 | 0.391 | 1.105 | 0.441 | 1.107 | 0.446 |
|  | tau<sub>MTL</sub> | 1.005 | 0.969 |  |  | 0.991 | 0.948 |
|  | HVa |  |  | 0.893 | 0.484 | 0.892 | 0.483 |
| Money | Aβ | 1.295 | 0.288 | 1.353 | 0.198 | 1.262 | 0.351 |
|  | tau<sub>MTL</sub> | 1.308 | 0.273 |  |  | 1.281 | 0.315 |
|  | HVa |  |  | 0.805 | 0.515 | 0.848 | 0.621 |
| Drive | Aβ | 1.138 | 0.442 | 1.185 | 0.297 | 1.136 | 0.451 |
|  | tau<sub>MTL</sub> | 1.170 | 0.336 |  |  | 1.168 | 0.346 |
|  | HVa |  |  | 0.960 | 0.848 | 0.988 | 0.955 |
| Work | Aβ | 1.259 | 0.387 | 1.379 | 0.206 | 1.241 | 0.422 |
|  | tau<sub>MTL</sub> | 1.540 | 0.064 |  |  | 1.510 | 0.080 |
|  | HVa |  |  | 0.747 | 0.373 | 0.798 | 0.492 |

**Note:** tauMTL: tau standardized uptake value ratios in medial temporal lobe, HVa: adjusted hippocampal volume, Aβ : Beta-Amyloid. Model 1 involves tauMTL + Aβ , model 2 involves only HVa + Aβ and model 3 involves three of them. Note: all models involve APOE4, Sex, Age and Education.

## Discussion

In a sample of cognitively unimpaired, Aβ positive individuals, we investigated the association of total and item-level Cognitive Function Index (CFI) scores with tau pathology, measured by the regional tau composite score of the medial temporal lobe (MTL), and neurodegeneration, measured by hippocampal volume. Our results indicate that higher tau_MTL_ levels and higher Aβ levels, at supra-threshold levels of Aβ positivity, were both associated with higher total CFI scores as reported by participants. The association between tau_MTL_ and total CFI scores reported by study partners was weaker and not significant. When investigating item-level CFI scores with AD biomarkers, we found that greater tau_MTL_ levels were associated with four items on the self-reported CFI and only one non-identical item on the study partner-reported CFI. The association between HVa and item-level CFI responses was weak and only significant for the noticeable memory decline item on both the self-report and study partner versions.

Previous studies demonstrated that Aβ+ individuals have higher self and study partner reported total CFI scores compared to Aβ-individuals [25]. It has also been shown that different items on CFI as reported by participants and their study partners were associated with higher amyloid burden in the A4 population [8]. In current study, we limited our sample to Aβ+ participants who were the primary target of the A4 trial. Even in this subgroup of participants, higher levels of Aβ were associated with higher total CFI scores as well as specific item-level responses. This suggests that the adverse effects of the underlying pathophysiology leading to amyloid accumulation may extend significantly beyond the levels typically associated with amyloid positivity.

Our results confirm previous findings that a higher self-reported total CFI score is associated with higher tau pathology in amyloid positive, cognitively unimpaired individuals [9]. Our research expands on these findings from different perspectives. Our main focus was in exploring the association of item-level responses to CFI with tau pathology. We found that distinct CFI items, whether reported by the participant or study partner, were associated with tau pathology. This may reflect the unique perspectives of participants and their study partners, suggesting that even in cognitively unimpaired and functionally independent individuals, both sources of information can effectively detect subtle cognitive changes.

Furthermore, we showed that the association of tau pathology with SCI at both global and item-level is largely independent of levels of amyloid pathology and neurodegeneration. This aligns with previous studies on SCI and AD pathology and may suggest multiple underlying pathways leading to SCI that do not necessarily interact to influence SCI endorsement [9, 26].

Another aim of our study was to explore the association between the CFI score and HVa, a marker of neurodegeneration. Given that neurodegeneration and hippocampal atrophy typically occur in the later clinical stages of AD, within the dynamic biomarker cascade [27], and since the A4 participants are all in the preclinical stage, it was not surprising to observe no significant association between HVa and total CFI score. Association between HVa and item-level CFI responses was weak and only significant for *challenges with the driving* item for participants, and *noticeable memory decline* observed by study partners. The hippocampus plays a crucial role in episodic memory, spatial memory and navigation [28] [29], so it is expected that impairment in these domains would correlate with earliest signs of hippocampal atrophy. Other studies have shown that subjective cognitive impairment predicts a faster rate of hippocampal atrophy [30, 31], so despite our weak association findings at cross-section, we expect to observe a similar pattern in the A4 study participants over the follow up period. This hypothesis can be evaluated in future studies with the release of the longitudinal phase of the A4 study.

Despite minor differences, overall consistency in endorsement patterns between participants and study partners validates the cognitive function items, indicating a shared perception of major cognitive challenges. However, items with the lowest endorsement rates for both sources, such as difficulties with appliances and financial tasks, did not show any association with AD biomarkers. These activities may be less frequently challenging for independent older adults and less critical for assessing SCI. Therefore, it may be worthwhile to explore the utility and sensitivity of a subset of the CFI questionnaire excluding these items as a marker for detecting cognitive changes in the earliest stages of the disease.

Our study has a few limitations. First, since CFI is a self-reported questionnaire, it is subject to recall bias, which may affect accuracy of responses by both participants and study partners. Inconsistent agreement between these sources may reflect differing perspectives and limited participant insight into specific items, highlighting the need for confirmation through multiple sources and during shorter time frames. Second, the majority of participants in the A4 study were white, limiting the generalizability of our findings to other racial and ethnic groups. Third, because the study design is cross-sectional, establishing causal relationships between different measures is not feasible.

Overall, our findings support that subjective reports of cognitive function can characterize early manifestations of cognitive impairment in preclinical AD trials. Longitudinal studies focusing on the association between subjective reporting of cognitive functioning and AD biomarkers in the context of AD trials is a future direction for this line of research.

## Data Availability

We have used A4 study which is publicly available on loni's website. https://ida.loni.usc.edu/login.jsp

## Acknowledgement

Current Research Funding: This research is supported in part by the National Institute of Health (NIA K23 AG063993; NIA-1R01AG080635-01A1); the Alzheimer’s Association (SG-24-988292) and Cure Alzheimer’s Fund.

Source of data and Funding: The A4 Study is a secondary prevention trial in preclinical Alzheimer’s disease, aiming to slow cognitive decline associated with brain amyloid accumulation in clinically normal older individuals. The A4 Study is funded by a public-private-philanthropic partnership, including funding from the National Institutes of Health-National Institute on Aging (U19 AG010483, U24AG057437, R01 AG063689), Eli Lilly and Company, Alzheimer’s Association, Accelerating Medicines Partnership, GHR Foundation, an anonymous foundation and additional private donors, with in-kind support from Avid and Cogstate. The companion observational Longitudinal Evaluation of Amyloid Risk and Neurodegeneration (LEARN) Study is funded by the Alzheimer’s Association (LEARN-15-338729) and GHR Foundation. The A4 and LEARN Studies are led by Dr. Reisa Sperling at Brigham and Women’s Hospital, Harvard Medical School and Dr. Paul Aisen at the Alzheimer’s Therapeutic Research Institute (ATRI), University of Southern California. The A4 and LEARN Studies are coordinated by ATRI at the University of Southern California, and the data are made available through the Laboratory for Neuro Imaging at the University of Southern California. The participants screening for the A4 Study provided permission to share their de-identified data in order to advance the quest to find a successful treatment for Alzheimer’s disease.

